# Admission photoplethysmography-based mortality prediction in hospitalized Ugandan children with suspected or confirmed infection: a feasibility study

**DOI:** 10.1101/2025.10.05.25337385

**Authors:** Mahan Rahimi, Matthew O. Wiens, Jerome Kabakyenga, Elias Kumbakumba, Nathan Kenya Mugisha, J. Mark Ansermino, Guy A. Dumont

**Affiliations:** School of Biomedical Engineering, University of British Columbia, Vancouver, Canada; BC Children’s Hospital Research Institute, Vancouver, British Columbia, Canada; Department of Anesthesiology, Pharmacology and Therapeutics, University of British Columbia, Vancouver, Canada; Institute for Global Health, BC Children’s and Women’s Hospital, Vancouver, British Columbia, Canada; Department of Community Health, Mbarara University of Science and Technology, Mbarara, Uganda; Holy Innocents Children’s Hospital, Mbarara, Uganda; Department of Paediatrics and Child Health, Mbarara University of Science and Technology, Mbarara, Uganda; Walimu, Kampala, Uganda; Department of Electrical and Computer Engineering, University of British Columbia, Vancouver, Canada

## Abstract

Sepsis remains a major cause of preventable pediatric hospital deaths in developing countries, with progress hindered by the lack of effective risk identification tools. Early detection of children at highest risk upon hospital admission is crucial for guiding clinical care and allocating resources, particularly in resource-limited settings. Photoplethysmography, which measures blood oxygen levels, also provides objective insight into cardiovascular alterations associated with sepsis. We conducted a secondary analysis of prospectively collected data from the Smart Discharges project, involving children under five years hospitalized with suspected or confirmed infection at six Ugandan hospitals, and developed models to predict all-cause in-hospital mortality across two age groups (0–6 and 6–60 months). Mortality was 7% in younger and 4.1% in older children. Machine learning models were trained on features extracted from one-minute photoplethysmograms collected at the time of admission. The best-performing model achieved mean values for the area under the receiver operating characteristic curve of 0.70 (95% CI: 0.62–0.76) in the younger cohort and 0.67 (95% CI: 0.56–0.73) in the older cohort, with corresponding values for the area under the precision–recall curve of 0.18 (95% CI: 0.12–0.27) and 0.14 (95% CI: 0.06–0.22), respectively. Calibration within risk strata was satisfactory (Brier scores 0.06 and 0.04), and decision curve analysis showed clinical utility. Notably, the models’ predictive capacity, although moderate, was achieved with a rapid and readily available objective measurement at admission, without the need for extended monitoring. While less accurate than most existing risk scores and not a substitute for clinical judgment, these simple admission-based models may help identify high-risk children and guide targeted interventions where sophisticated diagnostics are unavailable. External validation is needed before adoption.

**Author summary:** Sepsis poses a serious risk to children in hospitals with limited resources, and it can be difficult for healthcare workers to recognize which patients are in the most danger quickly. In our study, we investigated whether a simple fingertip sensor, commonly used in hospitals to measure oxygen levels, could help identify high-risk children upon arrival. We utilized data from Ugandan hospitals to develop computer-based tools that analyze signals from these sensors and support care decisions, eliminating the need for advanced equipment or laboratory tests. While our approach did not match the accuracy of the most advanced methods, it provided valuable information from a single, quick measurement at admission. These findings suggest that simple and accessible tools can still help staff make better decisions and prioritize children who require urgent care, even in settings with limited resources. We hope further work will refine these techniques and test their value in other hospitals and regions.

## 1. Introduction

The global burden of under-five mortality, especially from preventable causes, remains a pressing public health concern. Despite significant declines in recent decades, progress has been uneven, with sub-Saharan Africa accounting for more than half of all under-five deaths in 2023 [1–3]. This stark disparity reflects persistent systemic challenges, including limited healthcare access, inadequate infrastructure, and socioeconomic inequities [4–8]. Infectious diseases such as malaria, pneumonia, and diarrheal illnesses remain leading causes [1], often presenting with non-specific symptoms that can rapidly progress to sepsis—a life-threatening complication. In resource-limited healthcare settings, delays in triage and lack of diagnostic tools magnify these risks, underscoring the need for scalable approaches to improve child survival.

Rapid identification of children at high risk of mortality at admission is crucial, particularly in hospitals with overwhelming caseloads, minimal access to laboratory diagnostics, and where routine triage often relies on subjective assessments [9–11]. Timely and accurate risk stratification enables life-saving interventions and more efficient use of limited resources. Yet, most existing prediction models depend on detailed clinical or laboratory data, which are rarely available in low-resource hospitals [12]. This reveals an urgent need for simple, objective, and non-invasive methods that can be seamlessly implemented at the point of care, enabling accurate risk prediction upon admission.

Photoplethysmography (PPG) represents a promising modality for early and objective risk assessment in these environments [13]. As a non-invasive and affordable technology, PPG is most commonly acquired using portable devices such as pulse oximeters. It captures real-time relative changes in blood volume, enabling rapid data collection with minimal training and specialized clinical infrastructure [14]. This makes PPG particularly valuable in settings where high patient loads and limited resources preclude more complex diagnostics. PPG also provides reproducible physiological data, reducing the subjectivity associated with traditional assessments. Subtle waveform changes in PPG can reflect early physiological instability, and recent studies demonstrate that PPG-derived features are valuable in detecting and risk-stratifying sepsis, with increased variability and distinct pulse patterns identifying patients at higher risk of adverse outcomes [14–16]. This growing evidence supports the use of PPG as a frontline triage tool in low-resource settings.

In this context, we developed and evaluated mortality prediction models for children with suspected or confirmed infection at Ugandan hospitals, using only features derived from a one-minute admission PPG recording. Our objective was to assess the feasibility and utility of leveraging rapid, minimally burdensome physiological data for early risk stratification when comprehensive clinical or laboratory data are not available. We employed multiple machine learning algorithms on a broad set of PPG-derived features to assess predictive performance. While our models had moderate performance, comparable to some pragmatic clinical scores, they did not reach the accuracy of the most robust, multi-domain models. Nevertheless, an important advantage of this approach is that the application of a pulse oximeter requires no clinical assessment, whereas clinical prediction scores inherently demand clinician input and training. Thus, although PPG-based prediction may not replace the most accurate risk scores, it has the potential to meaningfully augment existing triage practices, addressing a critical gap where objective, easy-to-use decision support tools are otherwise lacking.

## 2. Materials and methods

### 2.1 Dataset description and approvals

Study data were accessed through the Pediatric Sepsis Data CoLaboratory [17], a global network for collaboration and data sharing to address pediatric sepsis morbidity and mortality [18]. Specifically, we used data from the initial observation phase (Phase-1) of the Smart Discharges project, a multi-center, prospective before-and-after study investigating the impact of tailored peri-discharge care on post-discharge mortality in Ugandan children <5 years hospitalized with suspected sepsis [19]. Ethical approval was granted by the Uganda National Institute of Science and Technology (HS 2207), the Mbarara University of Science and Technology Research Ethics Committee (No. 15/10-16, 27-Jan-2017), and the University of British Columbia–Children and Women’s Health Centre of British Columbia Research Ethics Board (H16-02679, 09-May-2017). Written informed consent was obtained from each child’s parent/guardian. Recruitment for Phase-1 included children aged 6–60 months (July 13, 2017–March 18, 2021) [20], and 0–6 months (January 15, 2018–June 30, 2023) [21]. An additional 0–6-month-old cohort was enrolled from March 31, 2020, to August 5, 2021, in response to COVID-19-related disruptions [22].

### 2.2 Population and setting

Eligible children were community admissions for suspected sepsis, defined by the treating team as confirmed or presumed infection. Prior work showed ∼90% of such patients met the International Pediatric Sepsis Consensus definition [23], which is specified as systemic inflammatory response syndrome with confirmed/suspected infection [24]. Exclusion criteria included residence outside the hospitals’ catchment area, admission for trauma, short-term observation (<24 h), and newborns admitted directly after birth without prior discharge to home. A newborn, or neonate, refers to a child in the first 28 days of life.

Participants were grouped into two cohorts: (i) <6 months, (ii) 6–60 months. Both were recruited at four Ugandan hospitals: Holy Innocents Children’s Hospital (Mbarara, southwest), Jinja Regional Referral Hospital (Jinja, east), Masaka Regional Referral Hospital (Masaka, central), and Mbarara Regional Referral Hospital (Mbarara, southwest). The <6 month cohort also included Uganda Martyrs Hospital (Ibanda, southwest) and Villa Maria Hospital (Masaka, central). During the study period, these public and faith-based facilities served both rural and urban populations of 30 districts (>8 million people, ∼1.4 million under-fives), representing the Ugandan pediatric population beyond the capital city of Kampala [25].

### 2.3 Data collection

A standardized protocol, with full details published previously [19,26], was used for all participants. At admission after consent, trained nurses collected clinical, social, and demographic data through caregiver interviews and clinical examination. Clinical data included vital signs; basic laboratory parameters (glucose, malaria rapid diagnostic test [RDT], human immunodeficiency virus [HIV] RDT, hematocrit, lactate); anthropometric measurements for nutritional assessment, converted to weight-for-height, weight-for-age, and height-for-age z-scores using the World Health Organization Child Growth Standards [27]; clinical signs and symptoms; co-morbidities; and medical history, including prior hospital admissions. Social and demographic variables included maternal and household characteristics. Participants were followed at 2, 4, and 6 months post-discharge; in- and out-of-hospital deaths and re-admissions were recorded.

### 2.4 PPG data preparation

Up to two one-minute PPG recordings per child were obtained with a LionsGate Technologies pulse oximeter and Android tablet running *Phone Oximeter* software [28]. Dual acquisitions increased the likelihood of capturing at least one high-quality waveform. The raw PPG consisted of infrared (940 nm) and red (660 nm) channels, sampled at 80 Hz; only the infrared data were analyzed due to their superior quality.

#### 2.4.1 Signal preprocessing

Recordings <30 s were excluded. All waveforms were temporally reversed to match standard PPG morphology. Preprocessing, adapted from Dall’Olio et al. [29], comprised several sequential steps. First, a 1-second centered moving average (CMA) was applied to suppress high-frequency noise while preserving low-frequency trends. This trend was subtracted from the original waveform, to remove baseline drift and isolate rapid fluctuations. To minimize amplitude variations and motion artifacts, the detrended signal was demodulated by constructing its analytic representation via the Hilbert transform. The instantaneous amplitude (envelope) was extracted, smoothed with a CMA, and used to normalize the detrended waveform, producing a denoised signal. A sample preprocessed PPG waveform is provided in S1 Appendix.

#### 2.4.2 Signal quality thresholding

Signal quality was assessed using the algorithm of Dall’Olio et al. [29], which yields a value reflecting both spurious noise events and variability in peak amplitudes. As lower values indicate better quality, this metric is referred to here as the *inadequacy score*. The score incorporates counts of extraneous local extrema and amplitude variance among detected peaks. For each child, only the highest-quality recording with a score <0.01 was kept.

Peak detection for this metric used a multi-scale, consensus-based approach: the CMA of each signal was computed using five window lengths (0.1, 0.5, 1.0, 1.5, and 2.0 s) and aligned with the waveform; signal segments above the CMA were identified at each scale; local maxima in these regions became candidate peaks; candidates from all scales were then compared, and only those present at every scale were retained. A 200 ms refractory period was imposed to avoid physiologically implausible detections [30]; if two peaks occurred within this interval, the higher-amplitude peak was kept.

#### 2.4.3 Feature extraction

PPG feature extraction was performed in three main steps: pulse segmentation, signal quality index (SQI) estimation, and feature quantification. A pulse was defined as the segment between the foot of one beat and the next. To improve fiducial point detection, signals were spline-interpolated, resampled from 80 Hz to 160 Hz, and normalized to a 0–1 range. Onsets were detected using a modified peak detection algorithm, with the signal inverted to identify minima; pulses were delineated between successive onsets.

SQI was computed using a slightly modified, correlation-based pulse-matching algorithm by Karlen et al. [31]. Forward and backward SQI values were obtained for each pulse, and the final SQI was taken as the maximum of the two. Only pulses with SQI >50 were retained. Recordings in which more than 70% of pulses fell below this threshold were excluded from further analysis.

Once high-quality pulses were identified, a comprehensive set of features was extracted. Derivatives of the PPG signal up to the fourth order were calculated to aid in locating fiducial points within each pulse. Zero-crossings of the fourth derivative were used to identify the *a*-, *b*-, *e*-, and *f*-peaks in the second-order derivative. Onset and systolic peaks on the PPG waveform (determined by the earlier peak detection method), along with the dicrotic notch and diastolic peaks (identified from the *e*- and *f*-peak positions), were then used to compute amplitude-, time interval-, and slope-based measures. Peak-to-peak intervals (PPIs) were computed from successive systolic peaks to generate a time series. From this series, time-, frequency-, and non-linear measures of pulse rate variability (PRV) were extracted, with frequency-domain analyses limited to recordings containing ≥30 s of continuous high-quality data. Oxygen saturation (*SpO_2_*) and perfusion index (pulsatile-to-non-pulsatile amplitude ratio) were exported directly from the pulse oximeter. For each feature, multiple pulse-level values were obtained per recording, with the median used for summarization unless otherwise stated. In total, 60 features were derived. After excluding those with pairwise correlation >0.8, 32 and 34 features remained for the 0–6-month and 6–60-month cohorts, respectively (Table 1). **S2 Appendix** includes the complete list of 60 extracted features and schematic illustrations of several examples.

**Table 1.** PPG-derived features and definitions grouped by extraction source.

| Feature, *unit* | Feature description |
| --- | --- |
| **Source: PPG** |  |
| crestTime, *ms* | Crest time (or risetime), time from onset to systolic peak |
| deltaTime, *ms* | Time delay between systolic and diastolic peaks |
| SysDiasTimeRatio | Ratio of time in systolic phase to time in diastolic phase |
| sysAmp, *a.u.* | Amplitude of systolic peak from onset |
| dicAmp, *a.u.* | Amplitude of dicrotic notch from onset |
| area1, *ms·a.u.* | Area bounded by times of onset and dicrotic notch |
| IPA | Inflection point area, ratio of area post- to pre-inflection point (dicrotic notch) |
| skewPulse | Pulse skewness, asymmetry of pulse waveform around the mean |
| kurtPulse | Pulse kurtosis, peakedness or flatness of pulse waveform |
| **Source: 2<sup>nd</sup> derivative of PPG** |  |
| abTime, *ms* | Time delay between a-peak and b-peak |
| a, *a.u.* | Amplitude of a-peak |
| b, *a.u.* | Amplitude of b-peak |
| b-a, *a.u.* | Amplitude difference between a-peak and b-peak |
| b/a | Ratio of b-peak amplitude to a-peak amplitude |
| abSlope, *a.u./ms* | Slope of straight line between a-peak and b-peak |
| **Source: PPI series** |  |
| Pulse rate, *bpm* | Number of pulses per minute |
| SDPP, *ms* | Standard deviation of PPIs |
| madPP, *ms* | Median absolute deviation of PPIs |
| tprPP | Turning point ratio* of PPIs |
| skewPP | Skewness of PPIs distribution |
| kurtPP† | Kurtosis of PPIs distribution |
| tprPPdiff | Turning point ratio of consecutive PPI differences |
| pNN20 (pNN50), % | Percentage of consecutive PPIs that differ by more than 20 ms (50 ms) |
| SD1/SD2 | Ratio of Poincaré plot standard deviation perpendicular to line of identity to that along line of identity |
| VLF, *ms<sup>2</sup>* | Absolute power of very-low-frequency band (0.0033–0.04 Hz) |
| LF, *ms<sup>2</sup>* | Absolute power of low-frequency band (0.04–0.2 Hz for newborns, 0.04–0.15 Hz for infants) |
| HF†, *ms<sup>2</sup>* | Absolute power of high-frequency band (0.20–2.00 Hz for newborns, 0.20–1.40 Hz for infants) |
| LF/HF | Ratio of LF to HF |
| LFn | Normalized power in low-frequency band (0.04–0.2 Hz for newborns, 0.04–0.15 Hz for infants) |
| entropyPP | Shannon entropy, unpredictability of PPIs |
| **Source: Pulse oximeter** |  |
| SpO<sub>2</sub>, % | Oxygen saturation |
| meanPERF | Mean of perfusion index |
| stdPERF | Standard deviation of perfusion index |

Abbreviations: bpm, beats per minute; ms: milliseconds.
\* The turning point ratio in a time series is a measure of the proportion of points where the direction of the series changes—specifically, where a value is a local maximum and/or minimum. Higher values indicate a more random series.
† Feature exclusive to the 6–60-month cohort. All other features were common to both the 0–6-month and 6–60-month cohorts.

### 2.5 Machine learning modelling

#### 2.5.1 Outcome definition

The primary outcome, peri-hospitalization mortality, was defined as all-cause death during hospitalization or within 48 hours of discharge. This included deaths shortly after leaving the hospital, such as among patients discharged against medical advice or who left without notice, as they were likely to have died had they remained in care. For brevity, the term mortality is used throughout the manuscript to refer to peri-hospitalization mortality.

#### 2.5.2 Model validation pipeline

Cohorts were analyzed separately: children <6 months and those aged 6– 60 months. In each cohort, survivors formed the majority class (label 0) and non-survivors comprised the minority class (label 1). Four machine learning classifiers—logistic regression (LR), random forest (RF), extreme gradient boosting (XGB), and multilayer perceptron (MLP)—were trained and compared. A support vector classifier was also evaluated but excluded due to poor fit. All models were implemented in Python 3.11.11 using scikit-learn 1.5.2.

Validation used nested stratified cross-validation (CV) with 5-fold outer and inner loops, repeated six times. In each outer iteration, data were split into training and test folds; all preprocessing, feature selection, and hyperparameter tuning confined to the training set in the inner loop. Hyperparameters were optimized via randomized search to maximize the area under the receiver operating characteristic curve (AUROC), with final performance assessed on the held-out outer test fold. Preprocessing comprised missing-value imputation by 5-nearest neighbours and normalization to a 0–1 range. The full hyperparameter search spaces for each model are detailed in **S3 Appendix**.

Feature selection occurred in the inner loop using recursive feature elimination (RFE) with LR as the estimator to identify the most informative predictors. Because the optimal number of features was unknown, multiple configurations were tested: in the 0– 6-month cohort, 18 models with RFE selecting 2–15 features (in single-feature increments), plus 20, 25, 30, and 32 features; in the 6–60-month cohort, the same approach was used, with a maximum of 34 features. After selecting the optimal feature set and hyperparameters, the model was retrained on the full outer training fold, calibrated using isotonic regression, and evaluated on the outer test fold. Reported results are averages across all nested CV repetitions.

As nested CV aims to produce unbiased performance estimates rather than a single fixed model, the selected feature sets varied between iterations. To identify the overall top predictors, Shapley Additive exPlanations (SHAP) methodology was applied across all folds [32]. Calculated SHAP values were averaged and weighted by the normalized frequency of feature selection, allowing identification of predictors that were both highly influential and consistently retained across model runs (Fig 1).

**Fig 1.**
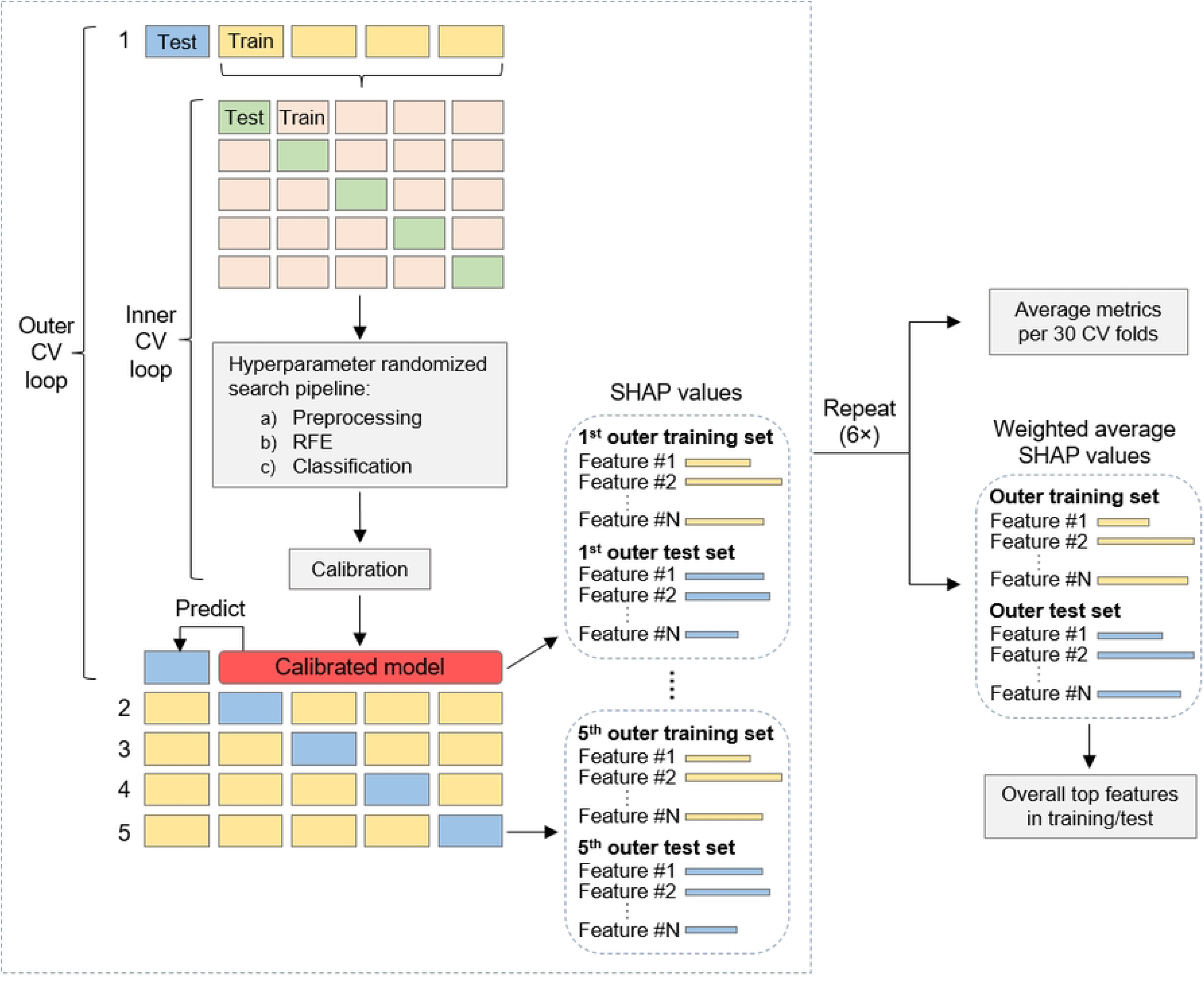
Overview of model validation pipeline. Model validation incorporated six repeats of 5-by-5 nested CV on observation data for hyperparameter tuning, feature selection, and model evaluation. The overall top features were identified using SHAP values averaged over all repetitions and weighted by the normalized frequency of feature selection.

#### 2.5.3 Performance metrics

Performance was evaluated in terms of discrimination, calibration, and clinical utility. Discrimination was measured by AUROC and the area under the precision–recall curve (AUPRC). Calibration was assessed visually with calibration plots and quantified using the Brier score. Clinical utility was examined with decision curve analysis, which estimates net benefit across threshold probabilities by comparing each model to the default strategies of treating all or none. Net benefit, defined in Equation 1, adjusts the proportion of true positives for the harm-to-benefit ratio of false positives, where *N* denotes total patients, *p_t_* is the threshold probability, and *p_t_/(1-p_t_)* represents the acceptable trade-off between harms and benefits [33].

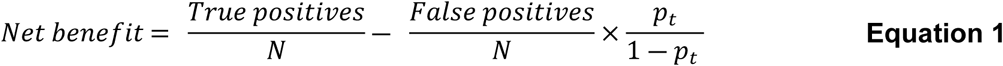

Additional metrics—specificity, positive predictive value (PPV), and negative predictive value (NPV)—were calculated to aid practical interpretation. Given the clinical context, the decision threshold was set to achieve a minimum sensitivity (recall) of 80%, ensuring at least 80% of high-risk patients were identified while limiting unnecessary interventions in low-risk cases.

## 3. Results

### 3.1 Study population

In the combined 0–6-month cohort during Phase-1 (including the COVID-19 enrollment), 2,707 hospitalizations were recorded (Fig 2). After excluding 274 cases with inadequate PPG recordings (low-quality, short, or absent data), 2,433 subjects remained: 2,263 (93%) survived and 170 (7%) died. Median length of stay (LoS), defined as days from admission to discharge or death for a single in-patient admission, was 5 days (interquartile range [IQR]: 3–6) for survivors and 3 days (IQR: 1–5) for non-survivors. In the 6–60-month cohort, 390 cases were excluded for the same reasons, leaving 3,449 subjects: 3,307 (95.9%) survived and 142 (4.1%) died. Median LoS was 4 days (IQR: 3– 6) for survivors and 2 days (IQR: 1–5) for non-survivors.

**Fig 2.**
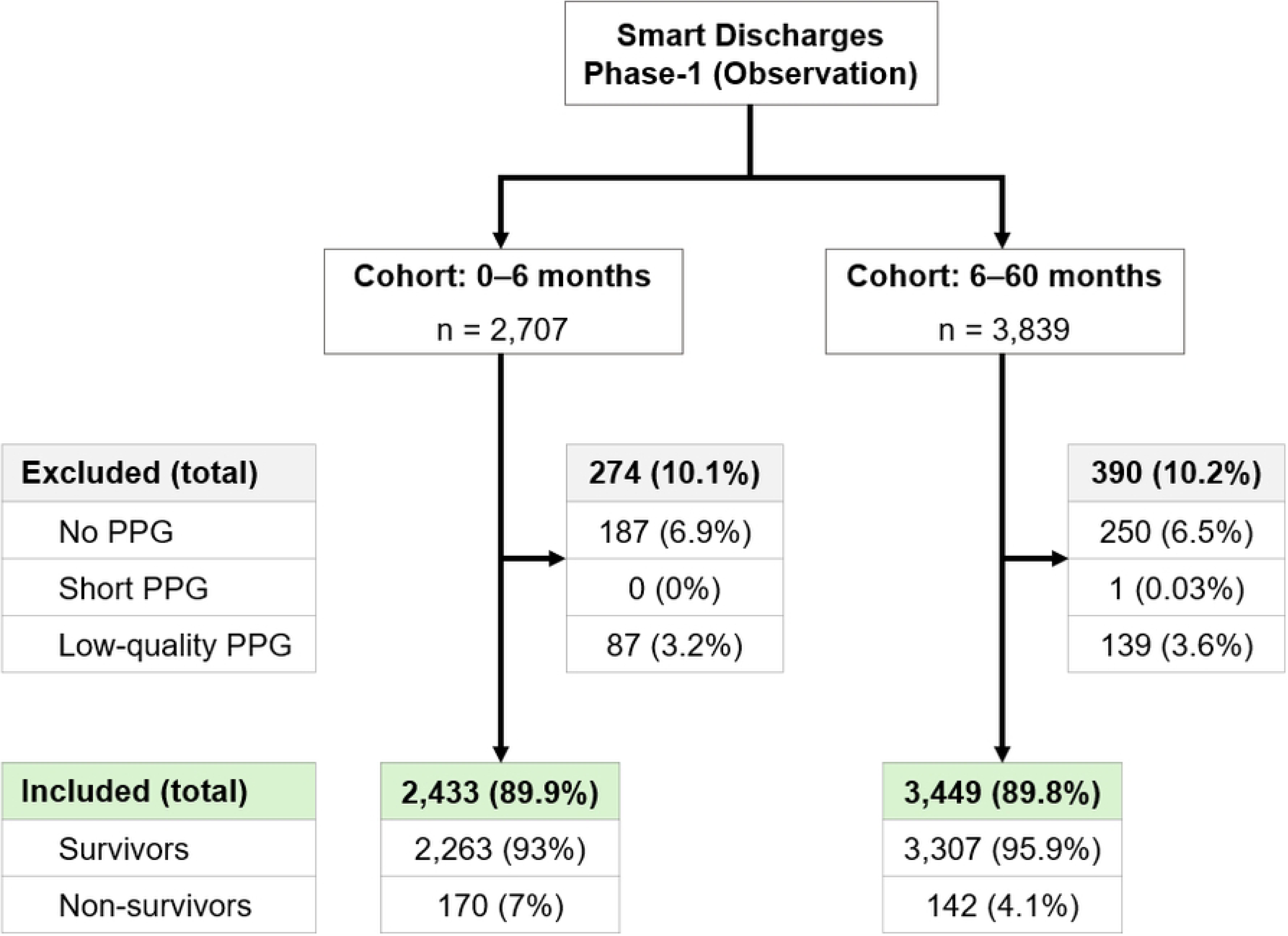
Population flow diagram, showing inclusions, exclusions, and outcomes for each age cohort.

Table 2 presents key baseline and admission features in the study population. Unless otherwise specified, numerical variables with multiple measurements per subject are reported as the median across all available measurements. Missing data were minimal, with no samples (rows) or features (columns) exceeding the predefined exclusion thresholds of 75% and 30%, respectively. Across both age groups, ∼55% of participants were male. In the 0–6-month cohort, median age was 1.5 months (IQR: 0.4– 3.6), and 13.6% had severe malnutrition (weight-for-age z-score < –3). In the 6–60-month cohort, median age was 16.6 months (IQR: 10.8–28), with severe malnutrition in 14.5% of participants.

**Table 2.** Baseline characteristics and admission PPG features in the study population, stratified by age cohort.

|  | 0–6-month cohort<br>(n = 2,433) |  | 6–60-month cohort<br>(n = 3,449) |  |
| --- | --- | --- | --- | --- |
| Variable | n (%) or<br>Median [Q1,Q3] | n Missing<br>(%) | n (%) or<br>Median [Q1,Q3] | n Missing<br>(%) |
| **A) Discharge characteristics** |  |  |  |  |
| Outcome |  | 0 (0%) |  | 0 (0%) |
| Survivors | 2,263 (93%) |  | 3,307 (95.9%) |  |
| Non-survivors | 170 (7%) |  | 142 (4.1%) |  |
| *Died in hospital* | 149 (6.1%) |  | 129 (3.7%) |  |
| *Died on discharge day* | 7 (0.3%) |  | 3 (0.1%) |  |
| *Died within 48hrs post-discharge* | 14 (0.6%) |  | 10 (0.3%) |  |
| Length of stay, days |  | 0 (0%) |  | 0 (0%) |
| Survivors | 5 [3,6] |  | 4 [3,6] |  |
| Non-survivors | 3 [1,5] |  | 2 [1,5] |  |
| **B) Demographics** |  |  |  |  |
| Sex, male | 1,357 (55.8%) | 0 (0%) | 1,907 (55.3%) | 0 (0%) |
| Age, months | 1.5 [0.4,3.6] | 0 (0%) | 16.6 [10.8,28] | 0 (0%) |

| C) Admission anthropometry |  |  |  |  |
| --- | --- | --- | --- | --- |
| Weight-for-age z-score | -0.7 [-2.0,0.2] | 1 (0.04%) | -1.2 [-2.3,-0.3] | 7 (0.2%) |
| < -3 | 331 (13.6%) |  | 500 (14.5%) |  |
| -3 to -2 | 284 (11.7%) |  | 547 (15.9%) |  |
| > -2 | 1,817 (74.7%) |  | 2,395 (69.4%) |  |
| D) PPG features |  |  |  |  |
| crestTime | 125 [106,150] | 0 (0%) | 106.3 [94,156.3] | 0 (0%) |
| deltaTime | 109 [106,112] | 0 (0%) | 106.3 [106,113] | 0 (0%) |
| SysDiasTimeRatio | 1.1 [0.9,1.3] | 0 (0%) | 0.9 [0.7,1.1] | 0 (0%) |
| sysAmp | 0.8 [0.7,0.9] | 0 (0%) | 0.8 [0.8,0.9] | 0 (0%) |
| dicAmp | 0.6 [0.5,0.6] | 0 (0%) | 0.6 [0.5,0.7] | 0 (0%) |
| area1 | 21.2 [19.6,23.1] | 0 (0%) | 19.5 [17.9,21.8] | 0 (0%) |
| IPA | 0.7 [0.5,0.8] | 0 (0%) | 0.8 [0.6,1.0] | 0 (0%) |
| skewPulse | 0.1 [0.0,0.2] | 0 (0%) | 0.2 [-0.0,0.4] | 0 (0%) |
| kurtPulse | -1.2 [-1.3,-1.2] | 0 (0%) | -1.2 [-1.3,-1.1] | 0 (0%) |
| abTime | 37.5 [37.5,43.8] | 0 (0%) | 37.5 [37.5,43.8] | 0 (0%) |
| a | 0.3 [0.2,0.5] | 0 (0%) | 0.5 [0.3,0.6] | 0 (0%) |
| b | -1.2 [-1.3,-1.2] | 0 (0%) | -1.2 [-1.3,-1.1] | 0 (0%) |
| b-a | -1.6 [-1.7,-1.5] | 0 (0%) | -1.7 [-1.8,-1.6] | 0 (0%) |
| b/a | -3.4 [-5.9,-2.3] | 0 (0%) | -2.5 [-3.6,-1.9] | 0 (0%) |
| abSlope | -0.04 [-0.04,-0.04] | 0 (0%) | -0.04 [-0.05,-0.04] | 0 (0%) |
| Pulse rate | 151 [136,165] | 0 (0%) | 145 [129,160] | 0 (0%) |
| SDPP | 11.5 [7.8,22.3] | 0 (0%) | 9.9 [6.8,17.2] | 0 (0%) |
| madPP | 6.2 [6.2,12.5] | 0 (0%) | 6.2 [6.2,6.2] | 0 (0%) |
| tprPP | 0.5 [0.4,0.5] | 0 (0%) | 0.4 [0.3,0.5] | 0 (0%) |
| skewPP | 0.2 [-0.1,0.6] | 0 (0%) | 0.1 [-0.2,0.5] | 0 (0%) |
| kurtPP | N/A |  | -0.1 [-0.4,1.2] | 0 (0%) |
| tprPPdiff | 0.3 [0.2,0.4] | 0 (0%) | 0.3 [0.2,0.4] | 0 (0%) |
| pNN20 | 3.8 [0.7,13.8] | 0 (0%) | 2.1 [0,10.7] | 0 (0%) |
| pNN50 | 0 [0,1.1] | 0 (0%) | 0 [0,0.8] | 0 (0%) |
| SD1/SD2 | 0.4 [0.3,0.5] | 0 (0%) | 0.4 [0.3,0.5] | 0 (0%) |
| VLF | 4.1 [1,16.4] | 251 (10.3%) | 3 [0.8,10.8] | 325 (9.4%) |
| LF | 14.4 [4.3,55.7] | 251 (10.3%) | 9.1 [3,26.3] | 325 (9.4%) |
| HF | N/A |  | 4.6 [1.6,15] | 325 (9.4%) |
| LF/HF | 1.7 [0.8,3.5] | 251 (10.3%) | 1.7 [0.8,3.3] | 325 (9.4%) |
| LFn | 0.6 [0.4,0.8] | 251 (10.3%) | 0.6 [0.4,0.8] | 325 (9.4%) |
| entropyPP | 4.9 [4.8,5] | 0 (0%) | 4.9 [4.8,5] | 0 (0%) |
| SpO<sub>2</sub> | 96 [92,99] | 0 (0%) | 97 [94,99] | 0 (0%) |
| meanPERF | 0.9 [0.6,1.5] | 0 (0%) | 1.3 [0.8,2.1] | 0 (0%) |
| stdPERF | 0.1 [0,0.1] | 0 (0%) | 0.1 [0,0.2] | 0 (0%) |

### 3.2 Model performance for mortality prediction

#### 3.2.1 Discrimination

In the 0–6-month cohort, all four models showed marked improvement in AUROC and AUPRC as the number of PPG features increased from 2 to ∼10, with the largest gains in the first few features (top row, Fig 3). AUROC then plateaued at ∼0.71 for RF and XGB, with LR and MLP reaching lower values. For AUPRC, LR and MLP peaked at ∼0.20–0.21, while RF and XGB leveled slightly lower (∼0.18–0.19). These patterns suggest that most predictive information was contained within the first 10 features, with later features adding minimal value.

**Fig 3.**
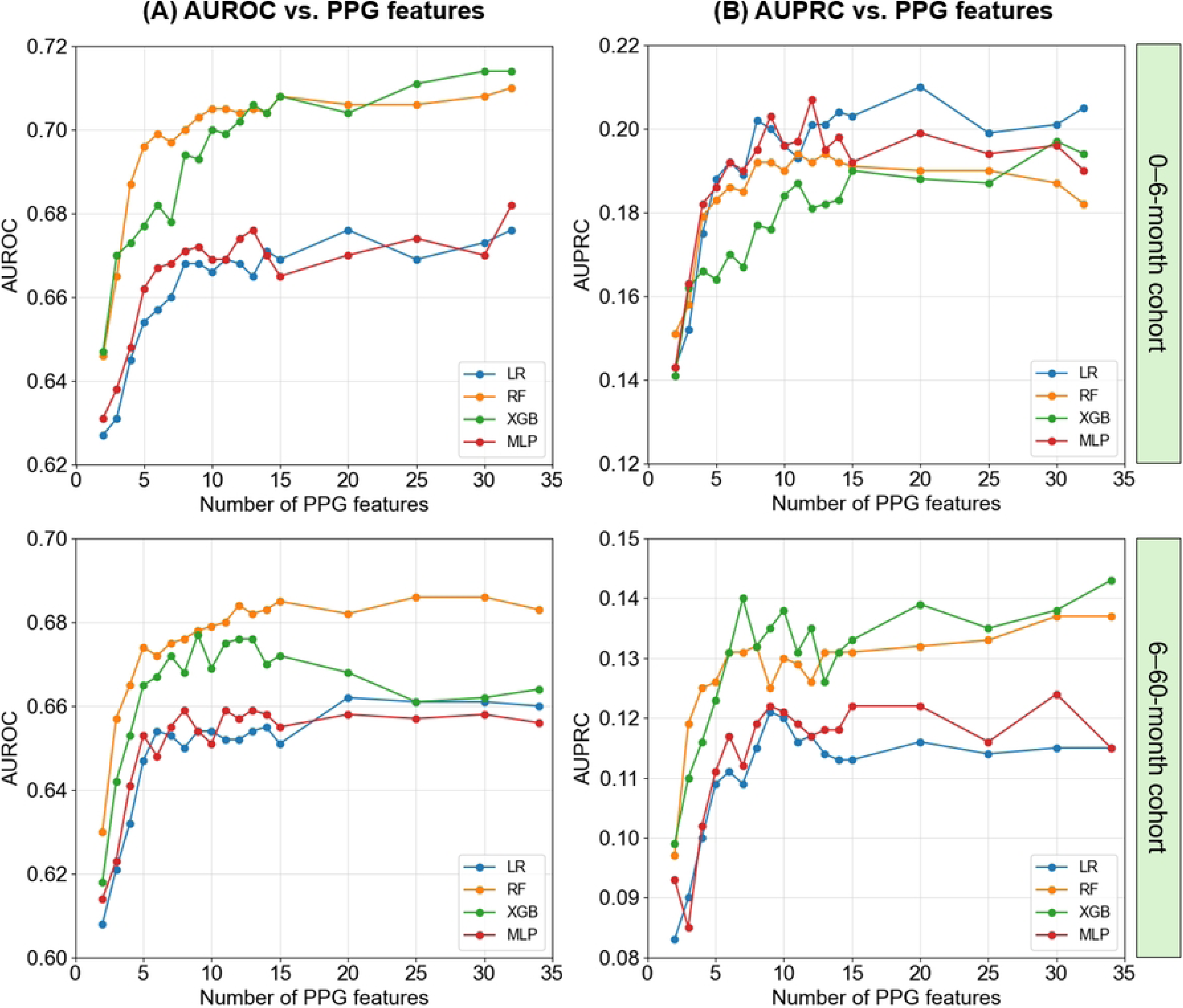
Discriminatory performance of models using different number of PPG features, stratified by age cohort. Mean values for AUROC (A) and AUPRC (B) across all CV test folds are shown for four models using varying numbers of PPG features to predict mortality. Results are shown separately for the 0–6 month and 6–60 month cohorts.

In the 6–60-month cohort, patterns were similar: performance improved substantially up to ∼10 features, then gains were marginal (bottom row, Fig 3). RF achieved the highest AUROC, closely followed by XGB, while LR and MLP plateaued at lower levels. For AUPRC, XGB reached the highest values (∼0.14), RF was close behind. LR and MLP had lower values. Again, most predictive value was captured within the first 10 features. Based on these trends, a 10-feature set was chosen for subsequent modeling, capturing nearly all attainable AUROC/AUPRC while preserving interpretability and avoiding unnecessary complexity. In the following sections, model names include the subscript “*_10_*” to denote 10-feature training.

In the 0–6-month cohort, across 30 CV iterations, LR*_10_*, RF*_10_*, XGB*_10_*, and MLP*_10_* achieved mean AUROCs of 0.67 (95% confidence interval [CI]: 0.59–0.75), 0.70 (95% CI: 0.62–0.78), 0.70 (95% CI: 0.62–0.76), and 0.67 (95% CI: 0.58–0.75), respectively, and mean AUPRCs of 0.20 (95% CI: 0.12–0.28), 0.19 (95% CI: 0.12–0.25), 0.18 (95% CI: 0.12–0.27), and 0.20 (95% CI: 0.12–0.29), respectively (Table 3). In the 6– 60-month cohort, discriminative performance was slightly lower, with AUROCs and AUPRCs as follows: LR*_10_*, 0.65 (95% CI: 0.56–0.73) and 0.12 (95% CI: 0.06–0.19); RF*_10_*, 0.68 (95% CI: 0.60–0.75) and 0.13 (95% CI: 0.07–0.22); XGB*_10_*, 0.67 (95% CI: 0.56–0.73) and 0.14 (95% CI: 0.06–0.22); MLP*_10_*, 0.65 (95% CI: 0.57–0.73) and 0.12 (95% CI: 0.07– 0.18). The mean receiver operating characteristic (ROC) and precision-recall curves of the four models in predicting mortality using 10 PPG features are shown in Fig 4.

**Table 3.** Performance summary and variables of 10-feature models for predicting mortality, stratified by age cohort.

|  | 0–6-month cohort |  |  |  | 6–60-month cohort |  |  |  |
| --- | --- | --- | --- | --- | --- | --- | --- | --- |
|  | LR<sub>10</sub> | RF<sub>10</sub> | XGB<sub>10</sub> | MLP<sub>10</sub> | LR<sub>10</sub> | RF<sub>10</sub> | XGB<sub>10</sub> | MLP<sub>10</sub> |
| **Metrics (95% CI)** |  |  |  |  |  |  |  |  |
| AUROC | 0.67<br>(0.59-0.75) | 0.70<br>(0.62-0.78) | 0.70<br>(0.62-0.76) | 0.67<br>(0.58-0.75) | 0.65<br>(0.56-0.73) | 0.68<br>(0.60-0.75) | 0.67<br>(0.56-0.73) | 0.65<br>(0.57-0.73) |
| AUPRC | 0.20<br>(0.12-0.28) | 0.19<br>(0.12-0.25) | 0.18<br>(0.12-0.27) | 0.20<br>(0.12-0.29) | 0.12<br>(0.06-0.19) | 0.13<br>(0.07-0.22) | 0.14<br>(0.06-0.22) | 0.12<br>(0.07-0.18) |
| Specificity | 0.35<br>(0.16-0.55) | 0.44<br>(0.29-0.61) | 0.42<br>(0.26-0.55) | 0.35<br>(0.15-0.60) | 0.34<br>(0.17-0.49) | 0.39<br>(0.20-0.56) | 0.38<br>(0.21-0.51) | 0.35<br>(0.17-0.54) |
| PPV (Precision) | 0.09<br>(0.07-0.12) | 0.10<br>(0.08-0.14) | 0.10<br>(0.08-0.12) | 0.09<br>(0.07-0.13) | 0.05<br>(0.04-0.07) | 0.06<br>(0.04-0.07) | 0.06<br>(0.04-0.07) | 0.05<br>(0.04-0.07) |
| NPV | 0.96<br>(0.93-0.98) | 0.97<br>(0.96-0.98) | 0.97<br>(0.95-0.98) | 0.96<br>(0.92-0.98) | 0.98<br>(0.96-0.99) | 0.98<br>(0.96-0.99) | 0.98<br>(0.97-0.99) | 0.98<br>(0.96-0.99) |
| Brier score | 0.06<br>(0.06-0.07) | 0.06<br>(0.06-0.07) | 0.06<br>(0.06-0.07) | 0.06<br>(0.06-0.07) | 0.04<br>(0.04-0.04) | 0.04<br>(0.04-0.04) | 0.04<br>(0.04-0.04) | 0.04<br>(0.04-0.04) |
| **PPG variables** |  |  |  |  |  |  |  |  |
| SpO<sub>2</sub> | ✓ | ✓ | ✓ | ✓ | ✓ | ✓ | ✓ | ✓ |
| Pulse rate | ✓ | ✓ | ✓ | ✓ | ✓ | ✓ | ✓ | ✓ |
| pNN20 | ✓ | ✓ | ✓ | ✓ | ✓ | ✓ | ✓ | ✓ |
| tprPP |  | ✓ | ✓ | ✓ | ✓ | ✓ | ✓ | ✓ |
| b | ✓ | ✓ | ✓ | ✓ | ✓ |  |  | ✓ |
| SD1/SD2 | ✓ | ✓ | ✓ | ✓ |  |  | ✓ | ✓ |
| LFn | ✓ |  |  |  | ✓ | ✓ | ✓ | ✓ |
| tprPPdiff | ✓ | ✓ | ✓ | ✓ |  |  |  |  |
| entropyPP | ✓ | ✓ | ✓ | ✓ |  |  |  |  |
| skewPP | ✓ | ✓ | ✓ | ✓ |  |  |  |  |
| sysAmp | ✓ |  | ✓ |  |  |  |  |  |
| crestTime |  | ✓ |  | ✓ |  |  |  |  |
| SysDiasTimeRatio |  |  |  |  | ✓ | ✓ | ✓ | ✓ |
| a |  |  |  |  | ✓ | ✓ | ✓ | ✓ |
| IPA |  |  |  |  |  | ✓ | ✓ |  |
| stdPERF |  |  |  |  |  | ✓ | ✓ |  |
| VLF |  |  |  |  |  | ✓ |  |  |
| dicAmp |  |  |  |  | ✓ |  |  | ✓ |
| skewPulse |  |  |  |  | ✓ |  |  |  |

The probability threshold was set to give a minimum sensitivity of 80%. The results represent average performance across 30 test folds obtained through repeated nested CV.

**Fig 4.**
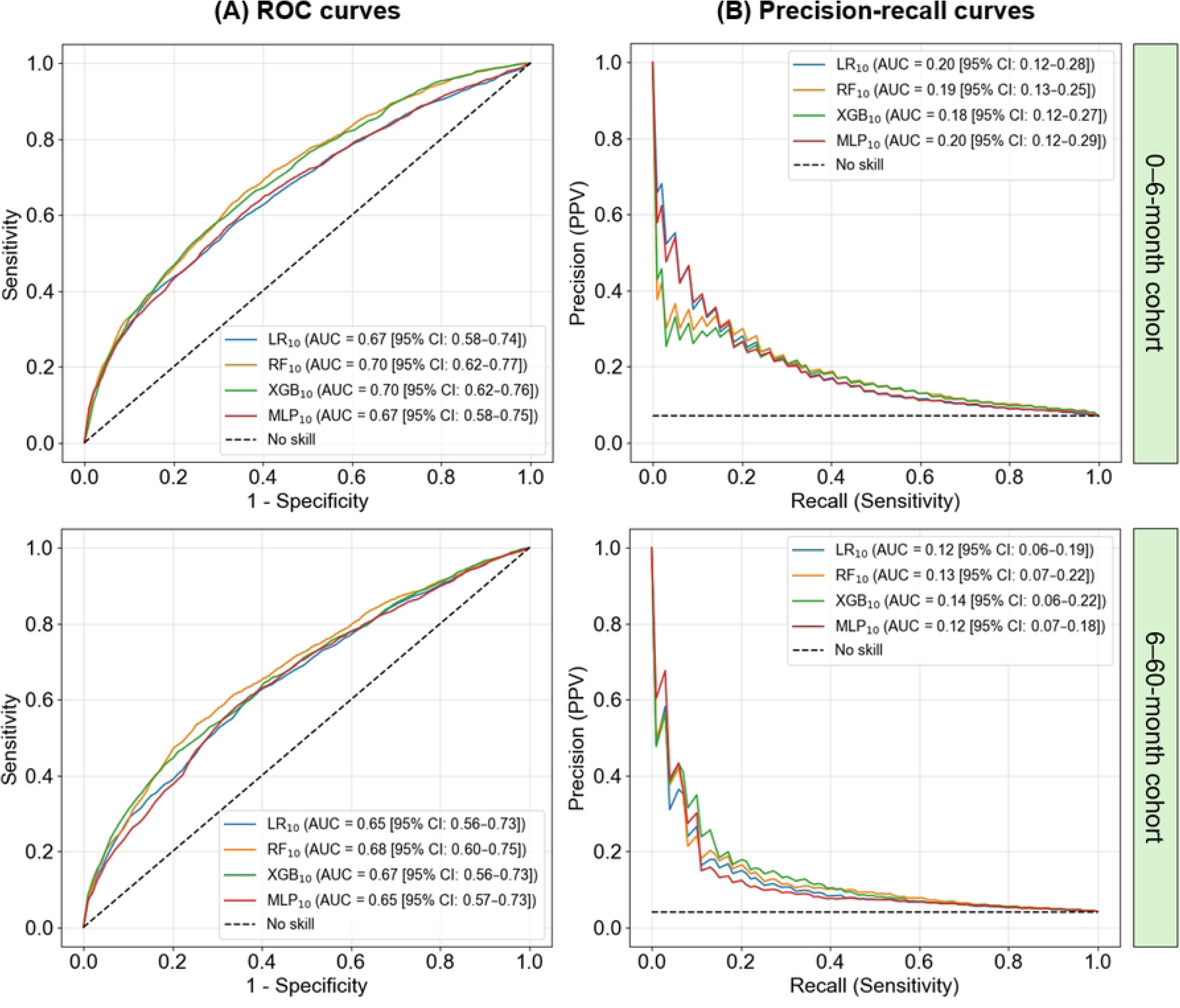
Discriminatory performance of 10-feature models obtained from test folds, stratified by age cohort. (A) ROC curves represent the mean plot across all CV test folds, illustrating the true positive rate (sensitivity) against the false positive rate (1-specificity). (B) Precision-recall curves show the mean plot across all CV test folds, illustrating the balance between precision (PPV) and recall (sensitivity). Results are shown separately for the 0–6 month and 6–60 month cohorts. Abbreviation: AUC, area under the curve.

In both age groups, RF*_10_* and XGB*_10_*achieved the highest discriminative performance, with mean AUROC values exceeding those of the other classifiers. The Friedman test confirmed statistically significant differences among classifiers (*p*-value<0.05), and subsequent Nemenyi post-hoc pairwise comparisons showed no significant difference between RF*_10_* and XGB*_10_* within either cohort (*p*-value≥0.05), indicating comparable AUROC performance. For AUPRC, results were less distinct: in the 0–6-month cohort, the Friedman test found no significant differences, whereas in the 6–60-month cohort, XGB*_10_* achieved significantly higher AUPRC than all other classifiers in pairwise comparisons, despite overall similarity in values. Pairwise Nemenyi post-hoc comparisons are reported in **S4 Appendix**.

With sensitivity fixed at 80%, average probability thresholds for RF*_10_* and XGB*_10_* were ∼0.05 (0–6-month) and ∼0.03 (6–60-month), respectively. At these thresholds, in the younger cohort, RF*_10_* achieved specificity = 0.44, PPV = 0.10, and NPV = 0.97, while XGB*_10_*yielded specificity = 0.42, PPV = 0.10, and NPV = 0.97. In the older cohort, both models had PPV = 0.06 and NPV = 0.98, with specificity of 0.39 for RF*_10_* and 0.38 for XGB*_10_*. NPV remained ≥0.97 across models and cohorts, indicating high reliability of negative predictions—an important property for ruling out mortality and safely deprioritizing immediate intervention for these patients. With NPV ≥0.97, fewer than 3% of children classified as low-risk would be expected to die.

#### 3.2.2 Calibration

Calibration plots (rows A and C, Fig 5) showed that all models were well-calibrated at lower predicted probabilities, with Brier scores of 0.06 for the 0–6-month cohort and 0.04 for the 6–60-month cohort. At higher probabilities—above ∼0.2–0.3 in the younger cohort and ∼0.1–0.2 in the older cohort—variability increased, and curves deviated from the diagonal, likely reflecting small sample sizes in these ranges and a tendency toward risk overestimation. Kernel density estimation (KDE) plots (rows B and D, Fig 5) displayed similar patterns across models and age groups. Predicted risks for survivors clustered near zero, whereas non-survivors showed broader distributions peaking at higher values. Substantial overlap occurred at very low risks (<0.1 in the younger cohort; <0.05 in the older), corresponding to both false-negatives (low-risk predictions for patients who died) and false-positives (high-risk predictions for survivors). Above ∼0.2 (younger cohort) and ∼0.1 (older cohort), non-survivor distributions became more distinct from those of survivors, indicating greater model discrimination in higher-risk ranges.

**Fig 5.**
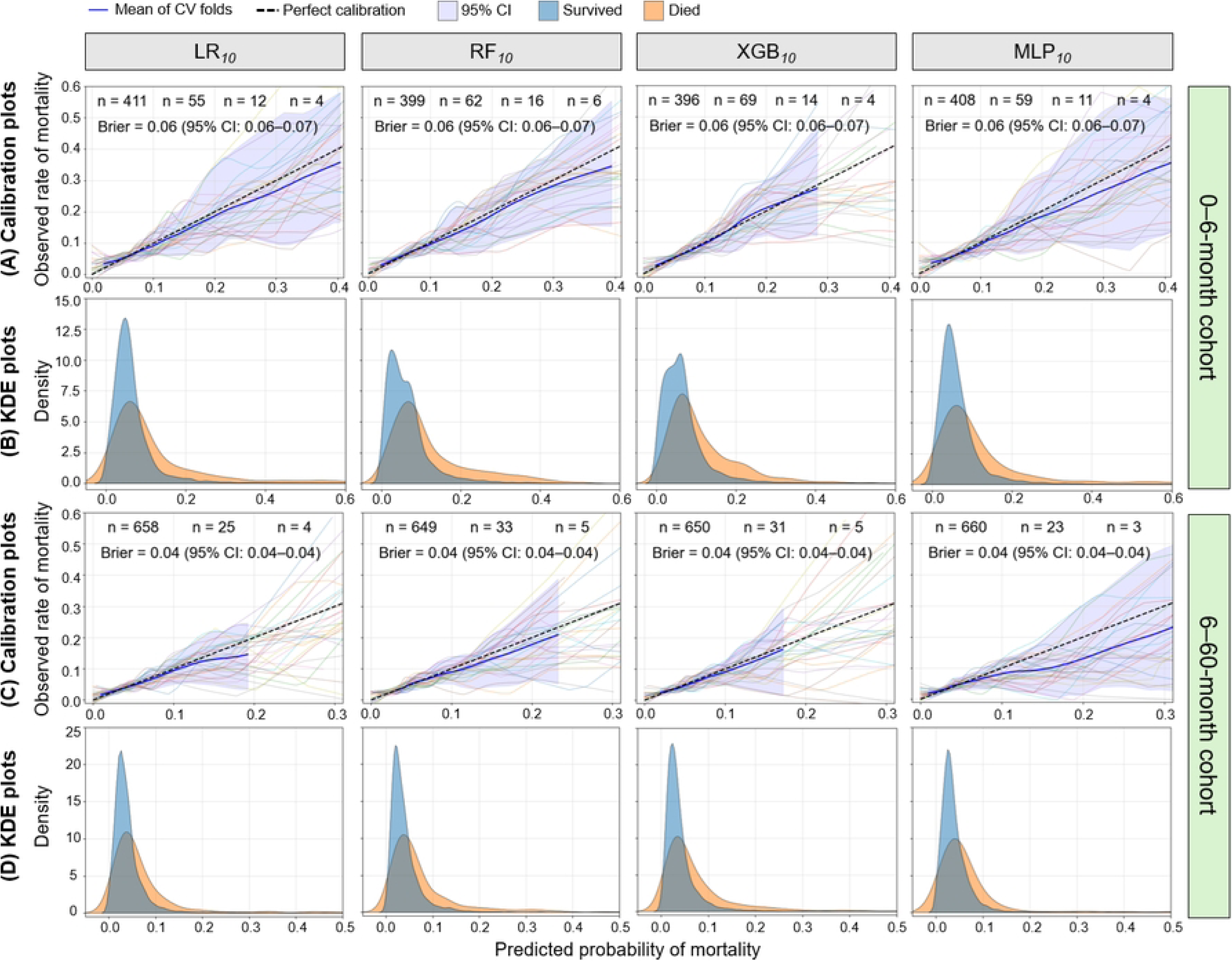
Calibration and KDE plots of 10-feature models obtained from test folds, stratified by age cohort. (A, C) Calibration plots compare each model’s predicted probabilities with the actual outcome. The solid blue line represents the mean curve across all CV test folds, while faint lines correspond to individual folds. (B, D) KDE plots illustrate the distribution of predicted probabilities for survivors and non-survivors. Results are shown separately for the 0–6-month and 6–60-month cohorts.

#### 3.2.3 Clinical utility

All four models showed greater net benefit for patient risk stratification than the default *treat-none-as-dying* strategy (assigning all patients to the negative class) across clinically relevant threshold probabilities in both age groups (Fig 6). In the 0–6-month cohort, net benefit exceeded that of *treat-none* for thresholds below ∼0.10; for the 6– 60-month cohort, this occurred for thresholds below ∼0.06. Compared with the *treat-all-as-dying* approach (assigning all patients to the positive class), all models outperformed within thresholds of ∼0.03–0.10 in the younger cohort and ∼0.02–0.06 in the older cohort. In the illustrated operating ranges, RF*_10_* and XGB*_10_*consistently provided slightly higher net benefit than the other models, with nearly overlapping curves, particularly in the older cohort. LR*_10_* and MLP*_10_* also achieved positive net benefit in the same threshold windows, showing fully overlapping curves in both cohorts. These findings indicate that all evaluated machine-learning models, and especially RF*_10_* and XGB*_10_*, offer meaningful clinical utility for triaging patients in both age groups compared with default strategies.

**Fig 6.**
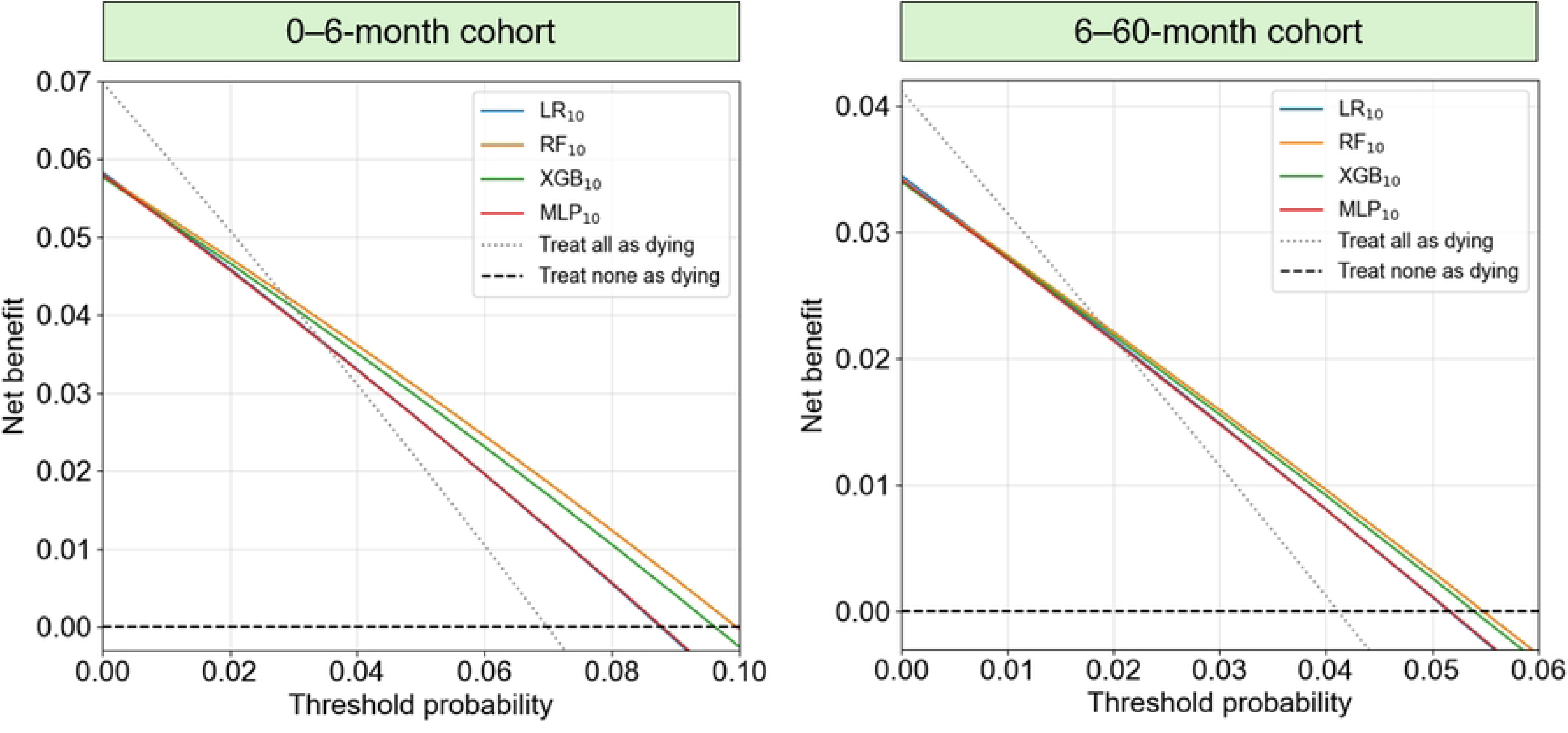
Decision curves of 10-feature models obtained from test folds, stratified by age cohort. The curves represent the mean net benefit across all CV test folds compared to the default clinical strategies of *treat-all* and *treat-none* at different threshold probabilities. Results are shown separately for the 0–6 month and 6–60 month cohorts.

At thresholds chosen to achieve 80% sensitivity in RF*_10_* and XGB*_10_* (0.05 for the 0– 6-month cohort and 0.03 for the 6–60-month cohort), the corresponding net benefit was approximately 0.03 and 0.016, respectively. In practical terms, this is equivalent to identifying 30 and 16 additional true positives per 1,000 patients compared with assuming all were negative, without increasing the number of false positives. At the same thresholds, the *treat-all* strategy produced net benefits of 0.021 for the younger group and 0.011 for the older group. Thus, RF*_10_* and XGB*_10_* improved net benefit by 0.009 and 0.005, corresponding to 171 fewer false positives per 1,000 patients in the 0–6-month cohort and 162 fewer in the 6–60-month cohort. This translates to reductions of approximately 17% and 16% in misclassification of low-risk patients, without increasing misclassification among those at high risk.

### 3.3 Predictors of mortality

In the 0–6-month cohort, a total of 12 features were identified as top mortality predictors across the four classifiers, with eight being consistently selected in every model and four displaying model-specific importance (Table 3). In the 6–60-month cohort, 14 features were identified, with seven consistently selected across all models, while the other seven demonstrated model-specific relevance. *SpO_2_* and *pulse rate*, direct measures of cardiorespiratory function, were selected in all models. There was also a clear dominance of PRV-related indices, including *pNN20*, *SD1/SD2*, *tprPP*, *tprPPdiff*, *LFn*, *VLF*, *entropyPP*, and *skewPP*. PRV has been widely recognized as a surrogate for heart rate variability (HRV) and a marker of autonomic nervous system function [34]. In sepsis, alterations in HRV and PRV have been linked to diminished physiological adaptability and impaired autonomic regulation in response to infection and systemic inflammation [35]. Decreased variability, lower entropy, and disruptions in pulse interval dynamics have independently predicted adverse outcomes in both pediatric and adult sepsis, supporting their use as early indicators of cardiovascular instability and poor prognosis.

Alongside PRV features, several amplitude- and morphology-based PPG features, including the *b*- and *a*-peak amplitudes, *sysAmp*, *IPA*, and *SysDiasTimeRatio*, were also identified as important predictors. These features have been shown to capture changes in vascular tone, arterial compliance, and microcirculatory function [15,29], which are characteristic alterations in sepsis. Specifically, attenuated systolic amplitude and distinct changes in the second-derivative waveform have previously been linked to endothelial injury and abnormal vasoregulation in septic patients.

As described earlier, both RF*_10_* and XGB*_10_* outperformed LR*_10_* and MLP*_10_*; however, XGB*_10_* was chosen for SHAP-based feature importance analysis due to its superior computational efficiency, and only its results are reported (Fig 7). See **S5 Appendix** for comparative feature importance plots from other models. In 0–6-month-olds, higher mortality risk was associated with lower *SpO_2_*, *b*, *skewPP*, *entropyPP*, and *tprPPdiff*, and with higher *SD1/SD2*, *pNN20*, *pulse rate*, *tprPP*, and *sysAmp*. *SD1/SD2* and *pNN20* were particularly robust, appearing in all 30 CV folds. The remaining predictors had selection frequencies ranging from 7 to 29 folds. In 6–60-month-olds, none of the top 10 predictors appeared in every fold, indicating less consistency across partitions. *pNN20* was the most frequent feature (28 folds). Lower *stdPERF*, *IPA*, *SpO_2_*, *a*, and *LFn*, and higher *pNN20*, *pulse rate*, *SysDiasTimeRatio*, *SD1/SD2*, and *tprPP* were linked to increased mortality risk.

**Fig 7.**
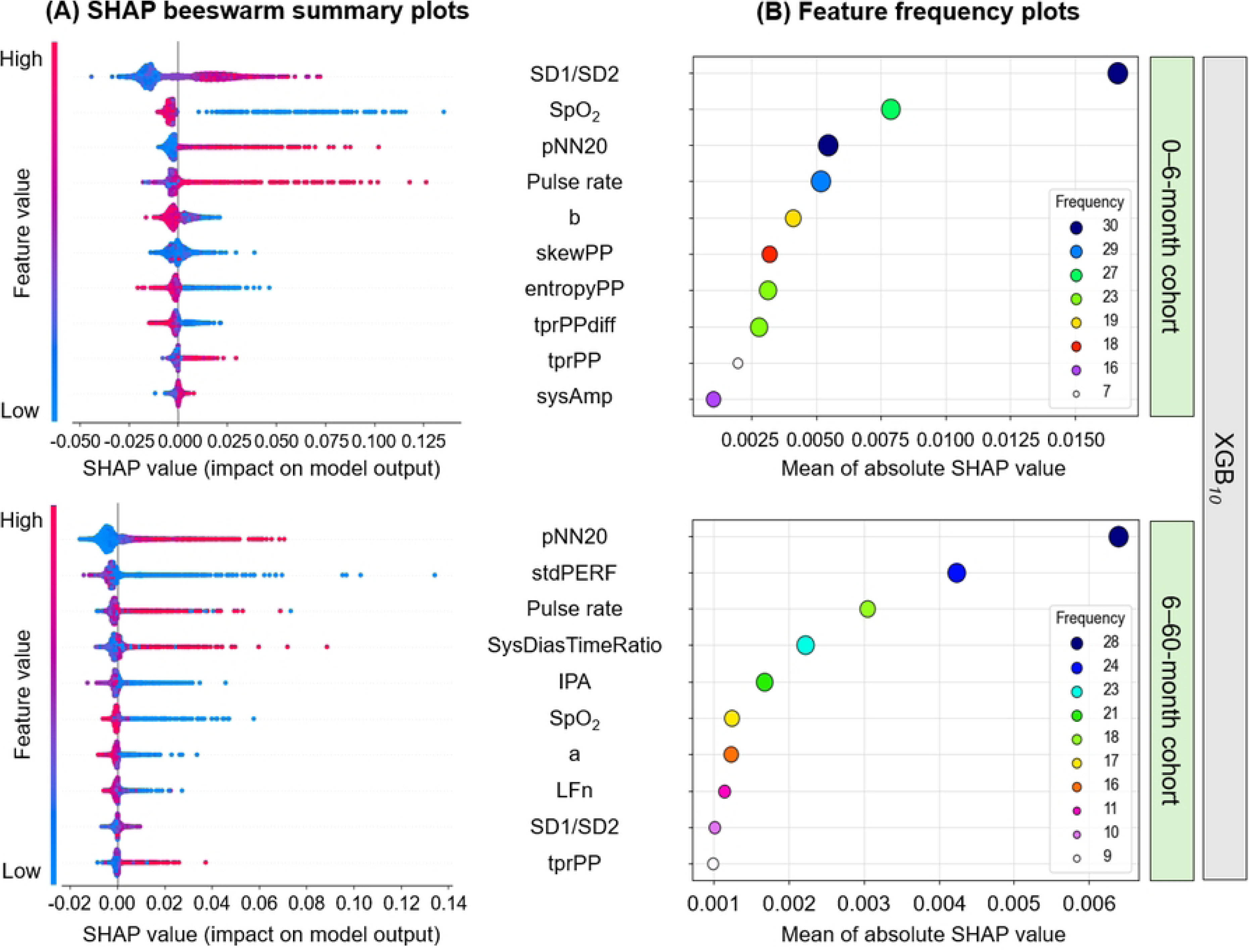
SHAP and frequency plots of XGB*_10_* obtained from test folds, stratified by age cohort. (A) The SHAP summary plot reflects the average results across all CV test folds. Each point represents an individual prediction, with SHAP values on the horizontal axis indicating the impact of features on the model’s output. Feature values are color-coded. Features are ranked by their mean absolute SHAP values, with the most important features appearing at the top. (B) The feature frequency plot shows the number of times each feature was selected across 30 CV folds, plotted against their mean absolute SHAP values. The size and color of the points represent selection frequency, with larger and darker points indicating higher frequencies. Results are shown separately for the 0–6 month and 6–60 month cohorts.

## 4. Discussion

In the rapidly evolving field of data-driven medicine, risk-differentiated care is increasingly recognized as a means to optimize treatment, particularly in resource-limited settings. Building on this principle, we developed and validated machine learning models based exclusively on one-minute admission PPG signals to predict hospital mortality among children under five admitted with suspected sepsis at multiple Ugandan sites. Through repeated nested cross-validation, we ensured robustness, and we used SHAP values to enhance interpretability. The purpose of these models is not to restrict access to care but to equip frontline healthcare workers with a rapid, objective prognostic tool, enabling timely interventions such as high-dependency unit placement or continuous monitoring of vulnerable children, tailored to each facility’s resources. To the best of our knowledge, no previous studies have explored models based solely on PPG features for pediatric hospital mortality in low- and middle-income countries (LMICs).

Our models demonstrated reliable calibration at lower predicted probabilities, and decision curve analysis indicated clinical utility across a spectrum of risk thresholds. Discriminatory performance (AUROC 0.67–0.70 in the younger cohort and 0.65–0.68 in the older cohort) fell within the mid-range of established tools, which report AUROCs from ∼0.55 to ∼0.85 [36–38]. While top clinical scores, such as the Lambaréné Organ Dysfunction Score (LODS) and the Signs of Inflammation in Children that Kill (SICK), demonstrate AUROCs up to 0.85, many commonly cited tools—including the modified Systemic Inflammatory Response Syndrome score (mSIRS) and the Pediatric Early Warning Score (PEWS)—report similar or only slightly superior discrimination to our PPG-based models. Importantly, many of the higher-performing traditional scores often depend on clinical, anthropometric, and laboratory variables—resources that are inconsistently available and subject to measurement variability in LMICs. In contrast, our approach utilized a rapid, non-invasive test that can be deployed by minimally trained staff, supporting objective and reproducible risk assessment at admission.

In our study, we trained four different models using small sets of PPG-derived variables. Extreme gradient boosting and random forest classifiers achieved the highest predictive performance, whereas the other models also showed competitive accuracy. Importantly, while there was considerable overlap in the features selected by different classifiers, some variability was also evident. This variability suggests a practical advantage: the availability of multiple models with comparable performance provides flexibility in deployment. Specifically, if certain features are unavailable, rendering one model (e.g., XGB) unusable, alternative models relying on different but overlapping feature sets can still be employed. This adaptability helps overcome challenges such as missing data at the point of care, ensuring a predictive tool remains available and effective.

We observed that different sets of features were selected across the two age groups, suggesting potential interactions between age and predictors. This age-related divergence may reflect underlying developmental differences in cardiovascular and autonomic regulation; however, further work is needed to determine whether certain PPG-derived indices hold unique prognostic value at specific ages. Another finding was that while both *SpO_2_*and *pulse rate* were identified among the top predictors, neither emerged as the top (i.e., most dominant) feature in either group. Given their established clinical association with illness severity, this result is surprising and merits further study. It may indicate that more subtle PPG-derived variability indices capture prognostic information beyond traditional vital signs, particularly in the context of sepsis.

Our study has several limitations which should be acknowledged. First, approximately 10% of subjects in both cohorts were excluded due to inadequate PPG signals, either failing to meet quality criteria or resulting from missing recordings. This data loss could introduce selection bias and may limit the generalizability of our findings; future efforts should focus on optimizing signal acquisition and preprocessing to minimize exclusions. Second, the outcome definition assumed that patients who left against medical advice or without notice were likely to have died had they remained hospitalized. While aligned with similar approaches in LMIC research, this assumption may not always be valid and could bias mortality estimates. Third, as this was a secondary analysis, the duration of available PPG recordings (one minute) was predetermined and could not be influenced. While ≥30 s segments allow extraction of most PRV features, reliable estimation of all indices generally requires ≥90 s [39]. Because not all recordings were analyzable, analyses were often restricted to 30 s, potentially reducing accuracy. Future studies should prioritize longer recordings to enable more comprehensive PRV analysis. Fourth, it is possible that predictive performance might be higher for outcomes occurring closer in time to admission (e.g., 48-hour mortality), than for peri-hospitalization mortality, an area that warrants future investigation. Fifth, the models were internally validated using cohorts from a single geographic region. Broader adoption will require robust external validation, tailored to the intended implementation context. Such validation should assess both geographic transportability—across different but plausibly similar populations—and methodological transportability, including varying data collection methods and conduct by independent investigators, and should ultimately include evaluation across time [40–42]. Finally, the actual clinical impact of our models remains unknown. Future prospective studies are needed to assess usability, acceptability, workflow integration, and real-world effects on patient outcomes and resource allocation.

In conclusion, although our models demonstrated only moderate performance, they may still provide practical value in settings with high patient-to-staff ratios and limited specialist availability, where structured risk stratification is often impractical. As a triage adjunct, the PPG-based models can serve as a rapid screening tool to identify higher-risk children, guiding expedited evaluation and resource allocation without replacing clinical judgment. The selection of PRV indices and other waveform-derived features as key predictors supports the physiological plausibility of this approach, given previous links between reduced variability and worse sepsis outcomes. By capturing early instability, the models could help focus limited resources on those most in need, ultimately improving pediatric outcomes in LMIC hospitals. Lastly, the moderate predictive performance likely reflects both the use of a single time-point measurement and reliance solely on physiological signals. To address this, we are now investigating ways to enhance accuracy by integrating additional minimally burdensome clinical or demographic variables and applying deep neural networks for automated feature extraction from raw PPG recordings.

## Data Availability

Study materials (dataset, data dictionary, and metadata) are publicly available through the Pediatric Sepsis Data CoLaboratory's (Sepsis CoLab) Dataverse: https://borealisdata.ca/dataverse/Pedi_SepsisCoLab. Due to the sensitive nature of clinical data and the potential risk for re-identification of research participants, the de-identified dataset is available through moderated access. Access to this data will be granted on a case-by-case basis following approval from the authors and the Data Governance Committees, and may be requested by contacting the CoLab Coordinator at.

## Acknowledgments

The authors extend their sincere gratitude to the caregivers/parents of the participants recruited, as well as to all current and former members of the Smart Discharges Research program: Matthew O. Wiens, Jeffrey N. Bone, Elias Kumbakumba, Stephen Businge, Abner Tagoola, Sheila Oyella Sherine, Emmanuel Byaruhanga, Edward Ssemwanga, Celestine Barigye, Jesca Nsungwa, Charles Olaro, J Mark Ansermino, Niranjan Kissoon, Joel Singer, Charles P. Larson, Pascal M. Lavoie, Dustin Dusmuir, Peter P Moschovis, Stefanie Novakowski, Clare Komugisha, Mellon Tayebwa, Douglas Mwesigwa, Nathan Kenya Mugisha, and Jerome Kabakyenga. Their dedication to data collection, administration, and logistical support has been invaluable.

## Supporting information sample

**S1 Appendix. Preprocessed PPG waveform.**

(DOCX)

**S2 Appendix. PPG-derived features and definitions.**

(DOCX)

**S3 Appendix. Hyperparameter search spaces for each model.**

(DOCX)

**S4 Appendix. Pairwise Nemenyi post-hoc comparisons.**

(DOCX)

**S5 Appendix. SHAP summary and frequency plots.**

(DOCX)

